# LUKB DT: A Web Tool for Quick and Efficient Identification of Disease Trajectories

**DOI:** 10.1101/2025.01.01.25319864

**Authors:** Xiangnan Li, Hui Zhang, Shuai Jiang, Baocai Gao, Zixin Hu

**Author notes:** Contributed equally (Xiangnan Li, Hui Zhang).

## Abstract

As the volume of electronic medical data grows, understanding disease progression and identifying risk factors have become central to public health research. Disease trajectories provide valuable insights into disease progression and risk factors. LUKB DT (LUKB Disease Trajectories) is a user-friendly web tool designed to facilitate disease trajectory analysis using electronic medical records (EMR), particularly the UK Biobank data. LUKB DT processes EMR data, including patient ID, disease diagnoses, corresponding timestamps, and other relevant variables, combining Cox regression, binomial tests, and conditional logistic regression to identify disease trajectories. This tool offers a quick and efficient way to prepare and analyze disease trajectories, contributing to the expanding field of disease trajectory research and providing valuable insights for risk factor identification and disease progression studies. Detailed deployment and usage can be found in the Supplementary Material.

LUKB DT is freely available at https://github.com/HaiGenBuShang/LUKB.

## Introduction

Large cohort studies have significantly advanced our understanding of complex diseases [1, 2]. The release of cohort data has fueled research across various fields, such as phenome-wide association studies [3], neuroscience [4], and epidemiology [5]. Electronic medical records (EMRs) offer a comprehensive approach to investigating the associations between diseases and other medical conditions [6, 7] by identifying so-called disease trajectories [5]. These trajectories have the potential to uncover genetic [8] and environmental risk factors [9], elucidate disease progression, aid in patient stratification, and provider further insights into disease mechanisms [10].

Disease trajectories represent the temporal order of disease occurrences, which can help identify key diseases [5, 10] that may lead to severe outcomes. While a disease trajectory browser has been developed for the Danish population [11], it primarily reflects the disease occurrence patterns within that specific cohort. As risk factors and disease progressions vary across populations, this tool may overlook important insights. With the increasing availability of large EMR datasets [1, 2], there is now an opportunity to identify disease trajectories and uncover unique risk factors and disease progressions in diverse populations.

The process of identifying disease trajectories typically involves four key steps (**Figure 1**), following data pre-filtering: (1) identifying diseases with increased risks associated with exposure to a disease or medical condition under investigation, (2) detecting disease pairs with temporal relationships, (3) confirming these disease pairs within datasets, and (4) concatenating disease pairs to form disease trajectories. To our knowledge, no public tool exists for this purpose. To address this gap, we have developed LUKB Disease Trajectory (LUKB DT), a web tool built on the R Shiny package and integrated into our UK Biobank data preparation platform, LUKB [12]. LUKB DT employs widely used strategies for identifying disease trajectories [10], streamlining the process with simple data input requirements and enabling researchers to generate meaningful insights efficiently. Furthermore, LUKB DT can be deployed locally, allowing researchers to analyze data securely on their own infrastructure without requiring extensive programming skills. In this paper, we present the LUKB DT tool and demonstrate its utility using UK Biobank EMR data.

**Figure 1.**
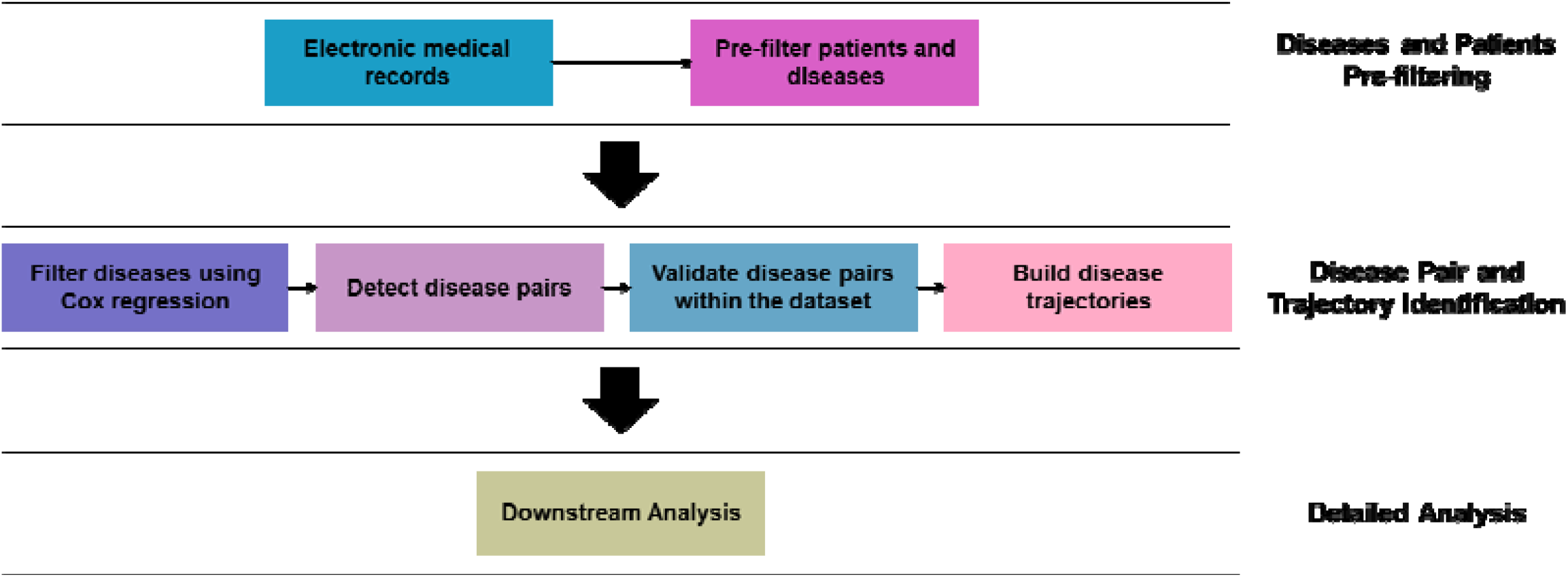
Workflow for constructing disease trajectories using electronic medical records. Pre-filtered EMR data undergoes four key steps to construct disease trajectories, which are subsequently used for detailed downstream analysis.

## Methods

### Tool design

The LUKB Disease Trajectory tool is an analysis component with four main parts (**Figure 2**) designed to identify disease trajectories, integrated into the LUKB platform. ‘Input data’ is designed for data input and setting analysis parameters for the entire process. ‘Disease filtering’ is responsible for identifying diseases with increased risks associated with the exposure under investigation. ‘Disease Pair Detection’ searches for candidate disease pairs with temporal occurrence orders. ‘Disease Pair Validation’ confirms the disease pairs detected in the previous step.

**Figure 2.**
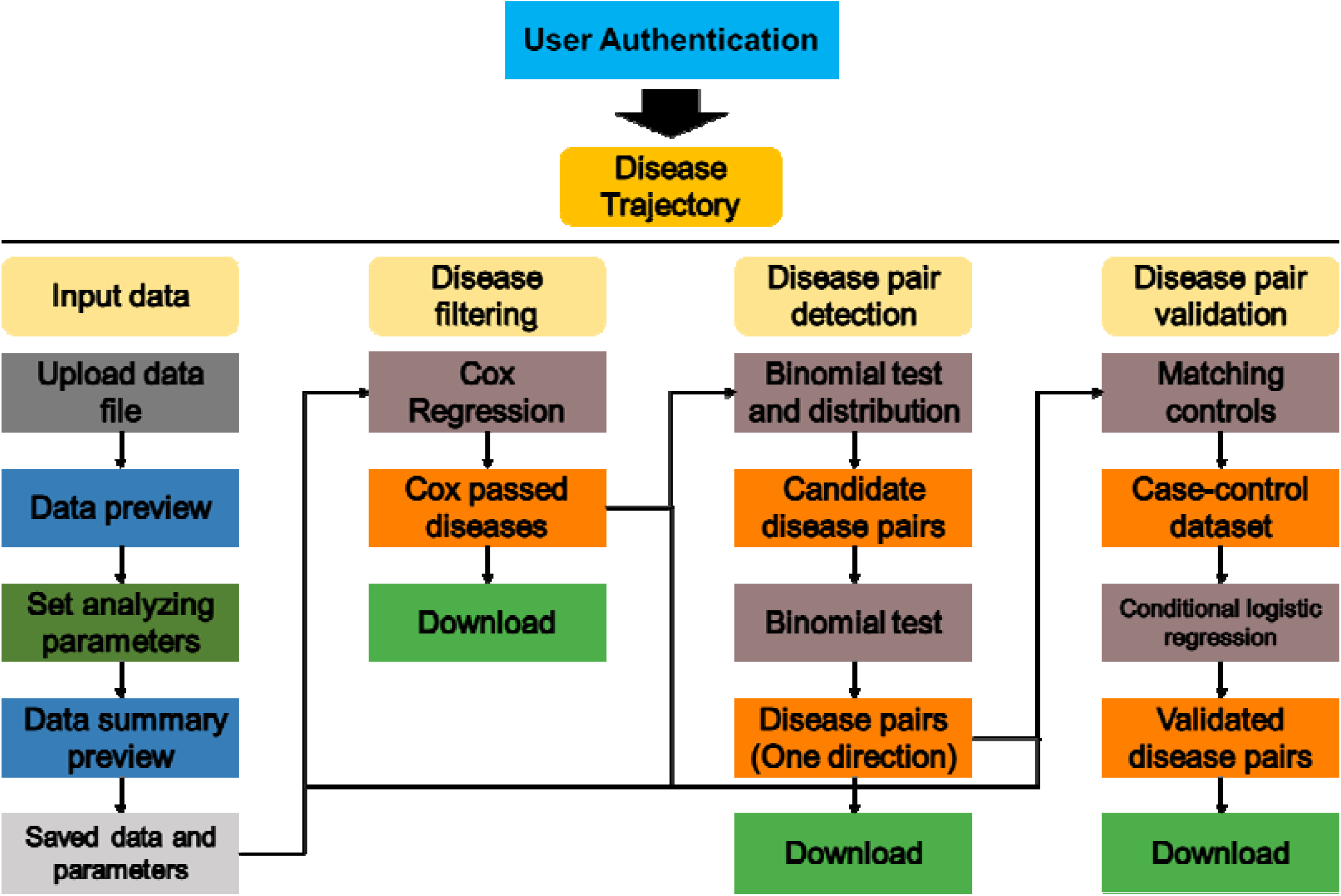
LUKB DT design. LUKB DT consists of four main parts.

#### Input data

In this part, the electronic medical records (EMR) data should be uploaded. The data must include at least participant IDs, the diagnosed diseases, and their corresponding diagnostic dates. Additionally, analysis parameters are set, including the exposure to be studied, minimum disease prevalence, study start and end dates, and adjustment variables. The follow-up period is defined as the time between the study start date and either the study end date or the diagnosis date. Details regarding these parameters are discussed in Section ‘Results and Discussion: LUKB DT Workflow’.

#### Disease filtering

To reduce the computational load of detecting and validating disease pairs with temporal occurrence orders, only diseases with an increased risk associated with the exposure under investigation are selected for further analysis. Cox regression analysis is conducted for each disease, with the exposure treated as a time-varying covariate. This step ensures that only diseases associated with the exposure are included in the next steps, minimizing the computational burden for subsequent analyses.

#### Disease pair detection

This part identifies candidate disease pairs with temporal occurrence orders using a Binomial test. Disease pairs with a length of 2 (D1 -> D2) were formed by pairing each disease from ‘Disease filtering’ part. Using the Binomial test, first, all disease pairs for which the p-value of the probability of getting disease D2 after D1 is significantly higher than the probability of getting D2 independent of D1 (p < 0.05) are selected. Next, the directionality of the disease pairs with palindrome directions is determined by testing which direction (D1 -> D2 or D2 -> D1) occurs more frequently among individuals. Statistical details are provided in Section ‘Methods Statistics: Binomial test to identify disease pairs and Binomial test to determine the directionality of disease pairs’.

#### Disease pair validation

In this final part, disease pairs that passed the last part are validated by confirming the direction of the disease pair using conditional logistic regression. First, a case-control dataset is generated based on the status of D2, with matching for adjustment variables. Then, a conditional logistic regression model is fitted, using D1 as the exposure and D2 as the endpoint. Disease pairs where the direction of the beta value for D1 is consistent with the direction in the Binomial test are considered true disease pairs with temporal occurrence orders. These validated disease pairs can then be used to construct disease trajectories.

### Statistics

#### Cox regression with time-varying exposure variable

The exposures under investigation may change over time in the electronic medical records (EMR) data. These exposures are treated as time-varying covariates in Cox regression analysis to maintain the proportional hazards assumption [13] and robustly estimate their effects on other diseases. After selecting a disease or medical condition as the exposure from the input EMR data, the start and end dates must be determined to define the follow-up period. The effects of the exposure on other diseases that meet the minimum prevalence criteria are estimated using Cox regression with the exposure as a time-varying covariate, along with other adjusting variables. Diseases found to be associated with the exposure and exhibiting increased risks are selected for further analysis with the binomial test to construct disease trajectories. For a more detailed explanation of individual and disease inclusion criteria, please refer to the **Supplementary Material**.

#### Binomial test to identify disease pairs

For diseases associated with the exposure and exhibiting increased risks, binomial tests are performed for all disease pairs (D1 -> D2). Each disease pair is assessed to determine if the prevalence of individuals who first developed D1 and subsequently developed D2 is significantly higher than the prevalence of individuals who developed D2 in the overall population.

#### Binomial test to determine the directionality of disease pairs

For each disease pair (D1 -> D2), a binomial test is used to assess whether more individuals experience D1 -> D2 than D2 -> D1, thus determining the directionality. Specifically, it tests whether the probability of developing D2 after D1 is significantly greater than 0.5 among individuals who developed both diseases. Only individuals with distinct dates for D1 and D2 are included, as identical dates could create ambiguity in determining the temporal order. For each disease pair, only one direction (D1 -> D2 or D2 -> D1) is retained, while the reverse direction is discarded.

#### Case control dataset and conditional logistic regression

Conditional logistic regression is used to confirm the directionality of disease pairs (D1 -> D2) using case-control datasets. These datasets are generated based on the status of D2 and matched for variables such as sex and age, which are provided in the input EMR data. Disease pairs (D1 -> D2) with consistent directionality between the binomial test and conditional logistic regression results are used to construct disease trajectories.

#### Disease trajectories

Disease trajectories are constructed by concatenating all confirmed disease pairs. For example, a D1 -> D2 -> D3 trajectory is formed if both D1 -> D2 and D2 -> D3 are confirmed disease pairs.

#### Conditional risk

Conditional risk refers to the likelihood of an individual developing D2 after developing D1, compared to the likelihood of developing D2 regardless of D1 status. It is calculated as:

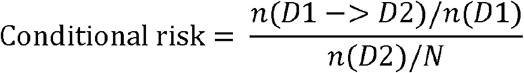

n(D1 -> D2) represents the number of individuals who developed D2 after D1, n(D1) represents the number of individuals who developed D1, n(D2) represents the number of individuals who developed D2, and N represents the total number of individuals included in the study.

## Results and discussion

### LUKB DT workflow

LUKB DT works from inputting EMR data to generating disease trajectories (**Figure 3**).

**Figure 3.**
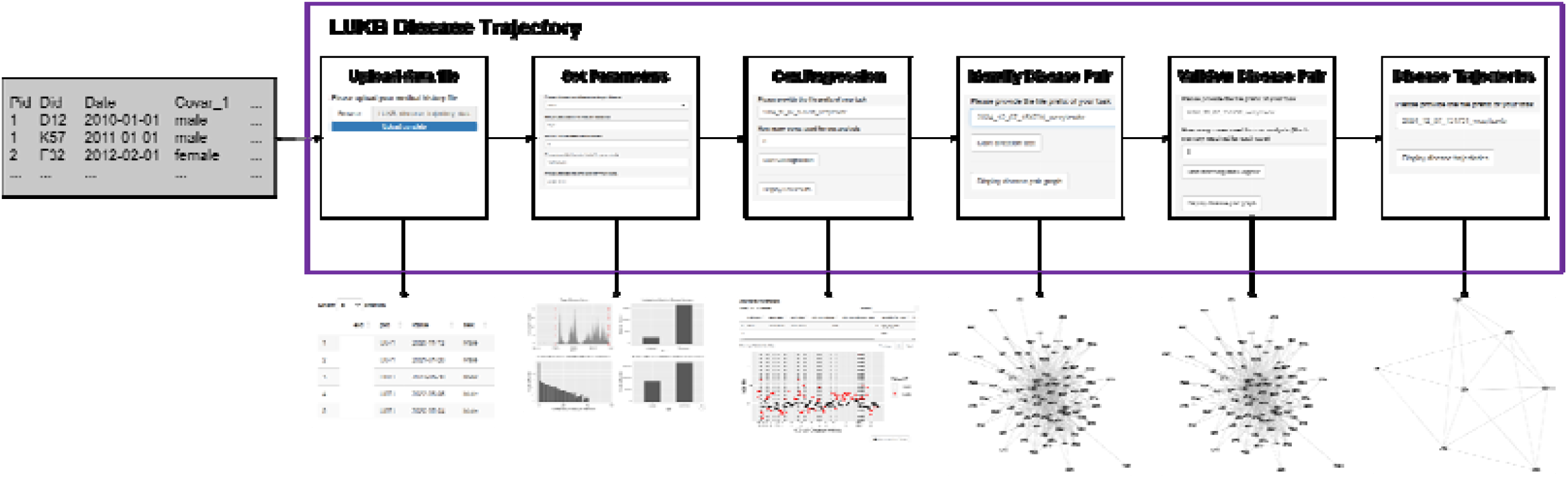
LUKB DT data processing workflow. LUKB DT requires EMR data with three essential columns in order: individual ID, diagnoses, and corresponding diagnosis dates. Additional covariates are optional. After uploading the data, users specify an exposure such as a disease or medical condition in the EMR data, and define analysis parameters. The tool then performs Cox regression, identifies disease pairs, and validates these pairs using conditional logistic regression, resulting in confirmed disease pairs. These confirmed pairs are concatenated to construct disease trajectories originating from the specified exposure.

#### Upload data and set analyzing parameters

LUKB DT begins by accepting EMR data with at least three essential columns in order: individual ID, ICD-10 diagnoses, and corresponding diagnosis dates. Users must select an exposure, either a disease or medical condition, from the input data for the analysis. The overall results will reflect the disease trajectories associated with this exposure. Based on the input data, LUKB DT filters individuals and diseases according to the specified analyzing parameters. The follow-up period is defined using start and end dates, with individuals who do not have any diseases during the follow-up period excluded from the analysis. Diseases with a prevalence below the specified threshold will also be excluded. A minimum gap between diseases is applied to exclude disease pairs at the individual level where the second disease occurs too soon after the first. Additionally, users can select adjusting variables for Cox regression and matching variables for conditional logistic regression to generate case-control datasets. The workflow provides a summary of the input data, including exposure cases per day, the number of included and excluded diseases, the number of individuals with different diseases, and the number of cases and controls.

#### Cox regression, disease pair detection and confirmation

Among the remaining steps, Cox regression is used to identify diseases significantly associated with the exposure, selecting those with a hazard ratio (HR) greater than 1 and an adjusted p-value < 0.05. These diseases are then analyzed to identify disease pairs. All disease pairs undergo binomial testing to assess whether the diseases in each pair occur in a temporal sequence. The conditional risk for each disease pair is calculated at this stage. Disease pairs with a Bonferroni-adjusted p-value < 0.05 and a conditional risk > 1 are selected for the directionality test. In this test, the temporal direction of each disease pair (D1 -> D2) is determined by testing whether more individuals experience the D1 -> D2 sequence than the reverse (D2 -> D1). Pairs that pass this test, with a Bonferroni-adjusted p-value < 0.05 and a positive coefficient for D1, are then further analyzed using conditional logistic regression to validate these disease pairs. A case-control dataset is created based on matching variables and the status of D2. Disease pairs that show statistical significance (Bonferroni-adjusted p-value < 0.05) and a positive coefficient for D1 are confirmed as valid disease pairs. The analysis allows the use of multiple computational cores for Cox regression and validation of disease pairs.

#### Disease trajectories

Once disease pairs are confirmed, they are concatenated to form disease trajectories, such as D1 -> D2 -> D3, based on the validated temporal associations.

### Case study: Disease trajectories of COVID-19 presenting by the UK Biobank ICD-10 diagnosis

The disease trajectories for depression, dementia, and breast cancer have been widely studied, with comprehensive assessments identifying the sequelae and pre-diseases for these conditions, offering insights for treatment and prevention [5, 6, 9]. The COVID-19 pandemic has caused a global health crisis, with millions infected worldwide. Subsequent diseases following COVID-19 infection have been extensively studied [14-16]. However, most studies have focused on specific diseases or disease classes, leaving a gap in comprehensively assessing the range of diseases that may follow COVID-19 infection. Using LUKB DT, we assessed the subsequent diseases that occur after a COVID-19 diagnosis. The results showed that COVID-19 primarily led to diseases and conditions related to SARS-CoV-2 infection, eventually resulting in other medical care as defined in ICD-10. In the meantime, these results reflect that the UK Biobank ICD-10 diagnosis data may not fully capture the disease trajectories related to COVID-19.

#### EMR dataset

We used the UK Biobank ICD-10 diagnosis data and corresponding diagnosis dates, alongside variables such as sex, year of birth, and date of inclusion (Field IDs: 41270, 41280, 31, 22200, and 53). ICD-10 codes were combined into 3-digit ICD-10 codes, and only the first date of diagnosis for each ICD-10 code was retained. For COVID-19, we specifically included the 4-digit ICD-10 code (U071) to differentiate it from its related ICD-10 codes “Emergency use of U07” (U071). Only individuals with a confirmed diagnosis were included in the analysis. In total, 6,166,668 medical records covering 446,995 individuals and 1,943 diseases/medical conditions were collected. Details of data preparation are provided in **Supplementary Material**. Among these individuals, 45.2% were male, and the ages ranged from 48.57 to 85.57 years on January 1, 2020.

#### Analyzing parameters for LUKB DT

We selected January 1, 2020, as the start date and October 31, 2022, as the end date, which represents the latest available follow-up date. The minimum prevalence and gap between two diseases were set to 0.005 and 0 days, respectively. A total of 183,386 individuals were included in the analysis, of which 8,411 had COVID-19, and 274 diseases were analyzed. Sex and age as of January 1, 2020, were used as adjusting variables for Cox regression, and sex and age groups as of January 1, 2020 were used as matching variables for conditional logistic regression. For the conditional logistic regression, one case was matched with two controls to generate the case-control dataset.

#### Identification of disease pairs

Cox regression identified 90 diseases significantly associated with COVID-19, distributed across 19 ICD-10 chapters (**Figure 4A**). Interestingly, no immune-related diseases, which have been reported to have an increased risk following COVID-19 [14-16], were significantly associated with the virus in this analysis. This may be due to the limitations of the ICD-10 diagnosis data, which primarily reflects diagnoses made within the UK’s primary care system and may not fully capture the entire spectrum of COVID-19-related diseases. These diseases formed 8,010 disease pairs, of which 2,410 exhibited a temporal association (Bonferroni-adjusted p-value < 0.05 and conditional risk > 1), and 527 passed the directionality test (Bonferroni-adjusted p-value < 0.05). All 527 disease pairs were confirmed by conditional logistic regression with a Bonferroni-adjusted p-value < 0.05 and a positive coefficient for the first disease.

**Figure 4.**
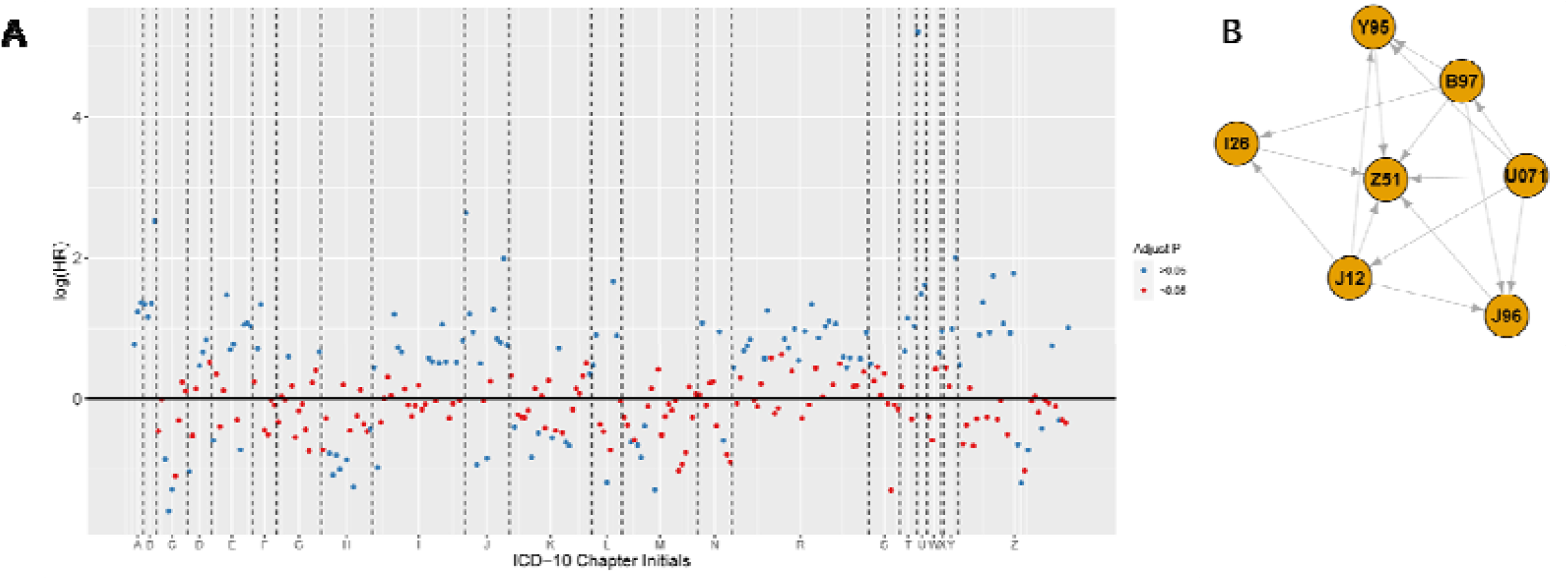
Diseases associated with COVID-19 and their formation into disease trajectories. A) Hazard ratio (HR) distribution of diseases meeting the minimum prevalence. Each point represents a disease, with red points indicating diseases significantly associated with COVID-19 based on Cox regression. The x-axis shows the initials of ICD-10 diagnoses, while the y-axis represents the log(HR). Diseases represented by red points above the horizontal black line were selected for constructing disease trajectories. B) Disease trajectories originating from COVID-19. Each vertex represents a disease, and arrows indicate the direction of disease pairs. The trajectories show that COVID-19 primarily led to SARS-CoV-2 infection-related diseases and eventually resulted in other medical care. U071: COVID-19 virus identified, B97: viral agents as the cause of diseases classified elsewhere, J12: viral pneumonia, not elsewhere classified, J96: respiratory failure, not elsewhere classified, Y95: nosocomial condition, Z51: other medical care.

#### Disease trajectories for COVID-19

The disease trajectories revealed that COVID-19 primarily led to SARS-CoV-2 infection-related diseases (ICD-10 codes: B97, J12, J96, Y95, Z51) and eventually resulted in other medical care (Z51) (**Figure 4B**). Specifically, diseases associated with viral agents (B97), viral pneumonia (J12), and respiratory failure (J96) could be direct consequences of COVID-19. While recent studies have linked COVID-19 to certain (auto)immune-related diseases [14-16], these conditions did not appear in our disease trajectories. This may be due to differences in diagnostic methods: those studies included SARS-CoV-2 test results, COVID-19-related hospital records, and rapid antigen tests, providing a more comprehensive view of COVID-19 cases. In contrast, the UK Biobank data primarily reflects COVID-19 cases diagnosed in hospital, which might underrepresent the COVID-19 status of individuals. Additionally, the risk of (auto)immune diseases may decrease over time following COVID-19 infection [15], which could explain the lack of temporal association in our analysis. Future studies on disease trajectories related to COVID-19 could benefit from incorporating EMR data that more accurately captures the COVID-19 status of individuals, such as combining ICD-10 diagnoses with primary care data from the UK Biobank, thereby enabling a more comprehensive understanding of the long-term health consequences.

### Benchmark

The performance of LUKB DT for constructing disease trajectories varies depending on the size of the medical records data, specifically the number of individuals and diseases or medical conditions included in the analysis. In the case study, the full calculation process took 55 minutes, using 8 computation cores for Cox and conditional logistic regression. The Cox regression took 11.4 minutes, the disease trajectory identification and confirmation took about 1 minute, and the conditional logistic regression took 42.3 minutes.

Benchmark tests for LUKB were conducted on an Intel(R) Core(TM) i9-9900K CPU @ 3.60 GHz, 32GB RAM server.

### Limitations and recommendations

One main limitation of LUKB DT is that it relies on processed medical records, such as ICD-10 diagnoses and their corresponding timestamps. Some diagnostic information may need to be manually extracted from the records [17, 18]. Another limitation is that the disease trajectories generated may not fully reflect the real trajectories experienced by patients [19]. This is because the disease trajectories are constructed by concatenating disease pairs that pass certain tests, rather than being directly derived from patient data.

Since the relationship between diseases in a pair is determined by their prevalence, it is crucial to use a comprehensive and representative dataset that includes all relevant individuals to ensure precise prevalence estimates. Therefore, we recommend that researchers include all individuals within the time interval to produce a more robust prevalence estimate for each disease. Additionally, we suggest that researchers view the disease trajectories presented by LUKB DT primarily as a tool for investigating risk factors or mechanisms for different diseases, rather than using them for patient clustering, due to the non-real nature of the disease trajectories.

## Conclusion

As electronic medical data continues to grow, its use in studying disease occurrence and mechanisms has become increasingly important. In this study, we introduced LUKB DT, a tool designed to identify disease trajectories using electronic medical records. LUKB DT streamlines disease trajectory analysis with easy data input and a user-friendly interface. Disease trajectories have become essential in studying risk factors, disease progression, and patient stratification. LUKB DT provides an efficient and rapid way to prepare disease trajectories, offering a valuable resource for future research in these areas.

## Data Availability

All data produced are available online at UK Biobank (https://www.ukbiobank.ac.uk/).

## Acknowledgements

We would like to thank the colleagues that participated in the development of LUKB, the Human Phenome Data Center at Fudan University and the Computing for the Future at Fudan (CFFF) platform for their assistance with data acquisition and processing. We would also like to thank all the authors who contributed to the R shiny or related packages and the funding agencies that supported this project.

## Authors’ contributions

X.L. conceived the idea, designed and developed the application, and drafted the manuscript. H.Z. developed the application, supervised the project, and revised the manuscript. J.S. and B.G. contributed to the development of the application. H.Z. supervised the project, drafted, and revised the manuscript.

