## Supplementary Material for "LUKB DT: A Web Tool for Quick and Efficient Identification of Disease Trajectories"

**Supplementary Materials**

**Individual and disease inclusion criteria**

Initially, LUKB DT excludes individuals based on the follow-up period and diseases based on minimum prevalence. Excluding individuals is intended to ensure the most reliable disease prevalence and the accuracy of the Cox regression. The exclusion of low-prevalence diseases is primarily aimed at maintaining statistical power. However, different criteria are applied for the Cox regression, binomial test for identifying temporal disease pairs, determining the directionality of disease pairs, and conditional logistic regression.

The process for filtering individuals and diseases in LUKB DT is described as follows (**Figure S1**): After excluding individuals without any diseases occurring within the follow-up period, Cox regression is used to identify diseases that form pairs for the binomial test, which then identifies disease pairs with temporal associations. The directionality of these pairs is subsequently tested, and those with confirmed directionality are used in conditional logistic regression.

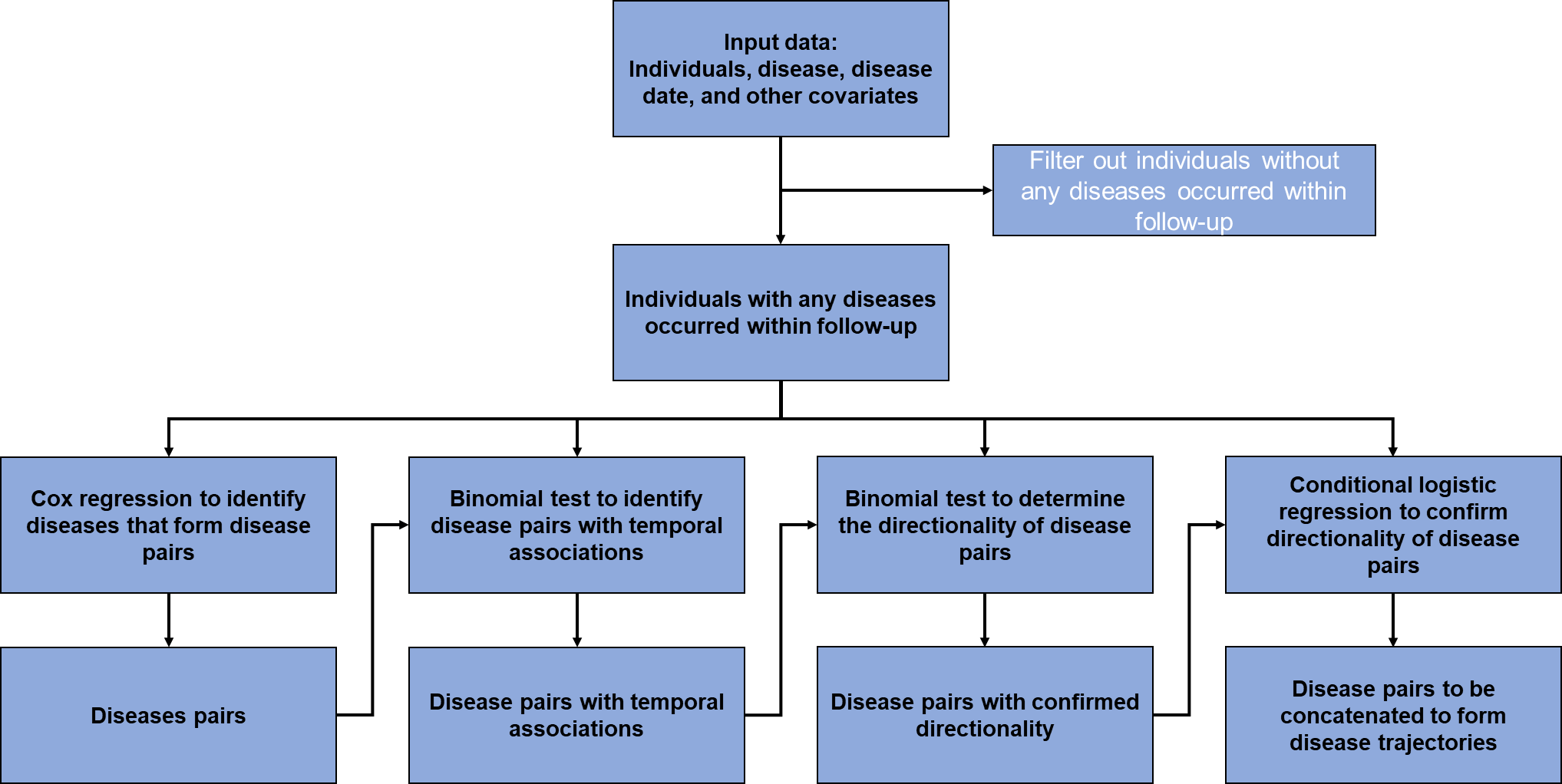

**Figure S1. Process for filtering individuals and individuals.**

***Cox regression***

For Cox regression, individuals with exposure occurring outside the follow-up period are excluded. Additionally, for each disease, individuals who developed that disease outside the follow-up period are excluded. If the prevalence of that disease meets the minimum threshold, Cox regression is performed with that disease as the response variable and exposure as the time-varying explanatory variable, adjusted for other covariates (**Figure S2**). Diseases significantly associated with exposure (Bonferroni-adjusted p-value < 0.05 and Hazard ratio > 1) are then used to form disease pairs.

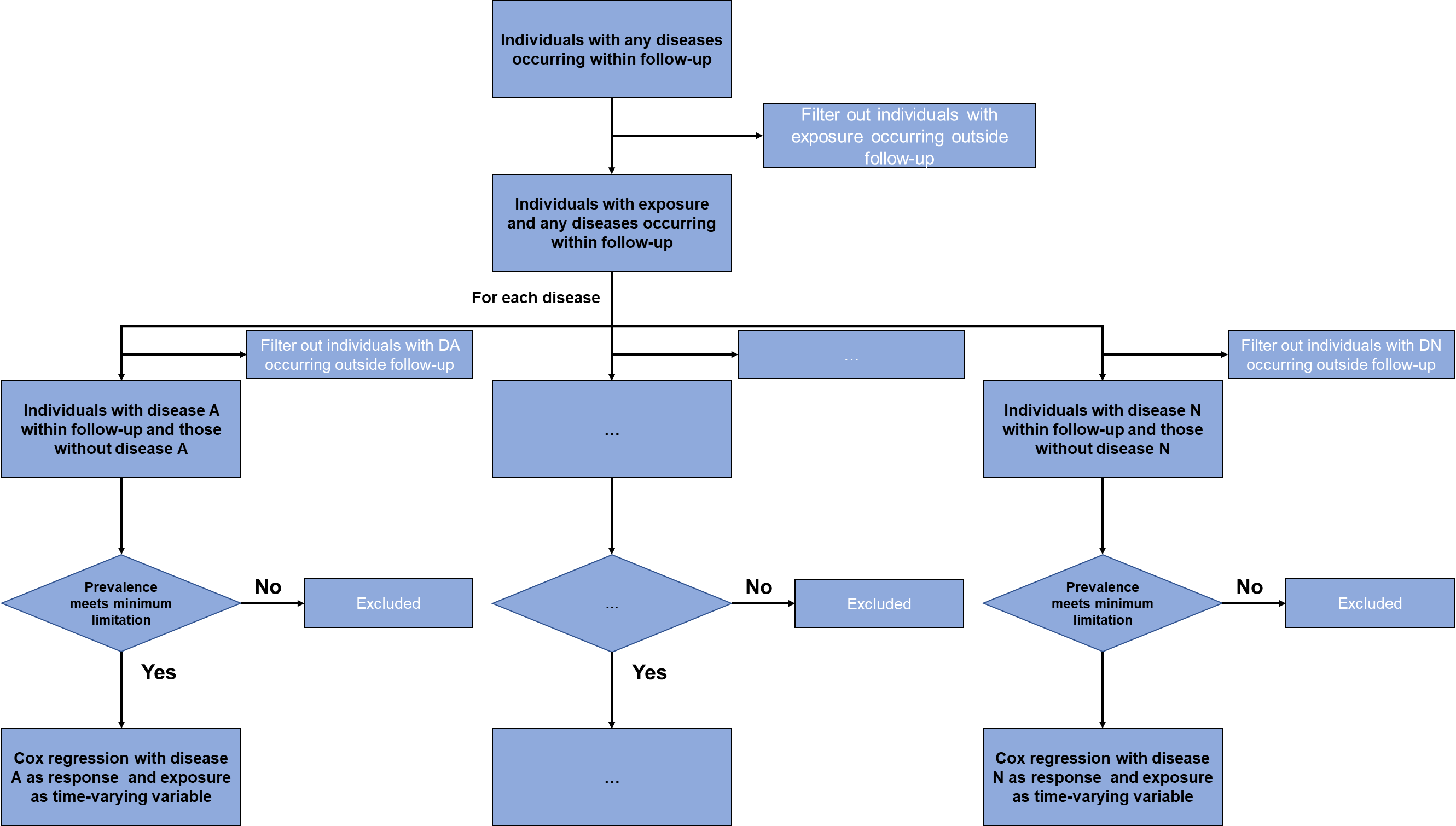

**Figure S2.** **Process for filtering individuals and diseases in Cox regression.**

***Binomial test to identify disease pairs with temporal associations***

For each disease pair (D1 -> D2), only individuals with D1 occurring within the follow-up period and D2 occurring before the end of the follow-up period are included. These individuals are counted as having D1. Individuals with D2 occurring before D1, with D2 occurring after D1 but within the minimum disease gap, or with D2 occurring within the follow-up period, are also included. These counts, along with the total number of included individuals, are used in the binomial test to identify temporal associations between disease pairs (**Figure S3**). Disease pairs with a temporal association are defined as those with a Bonferroni-adjusted p-value < 0.05 and a conditional risk > 1.

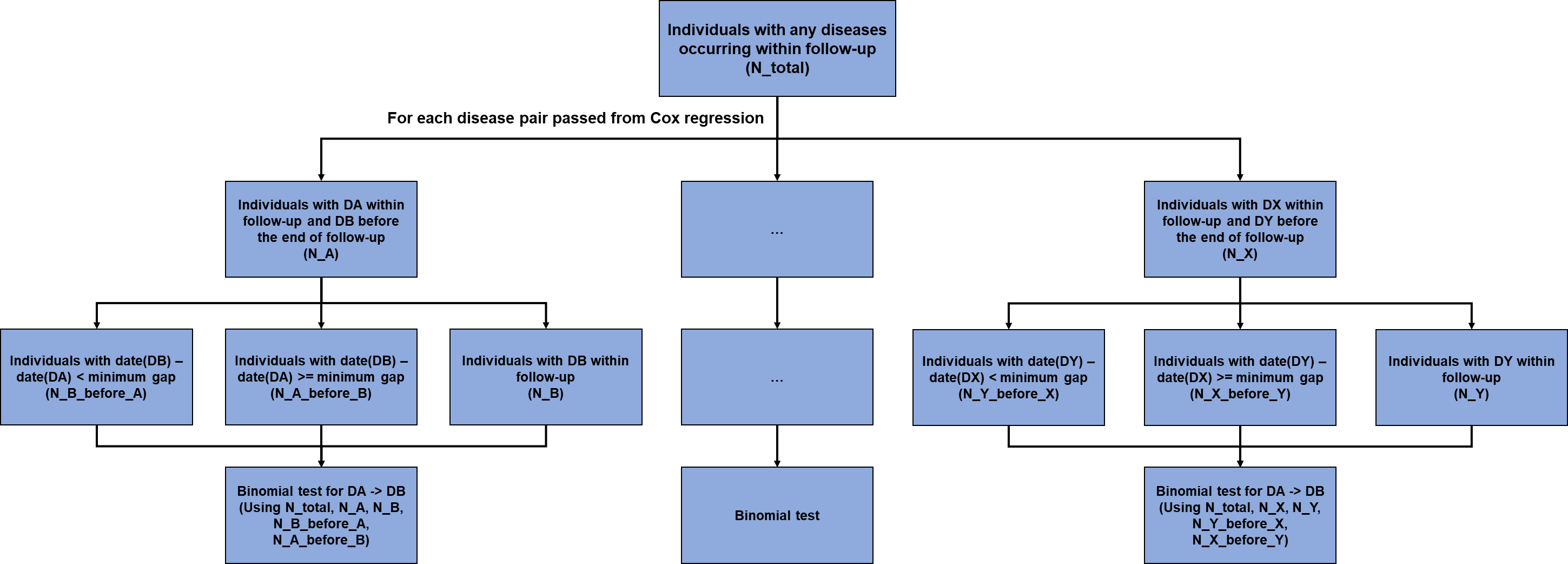

**Figure S3.** **Process for filtering individuals and diseases in the binomial test to identify temporal associations between disease pairs.**

***Binomial test to determine the directionality of disease pairs***

For disease pairs with identified temporal associations (D1 -> D2), individuals with both D1 and D2 occurring within the follow-up period are included. The numbers of individuals with D2 occurring before and after D1 are then counted. These counts are used in the binomial test to determine the directionality of the disease pairs (**Figure S4**). Disease pairs with confirmed directionality are those with a Bonferroni-adjusted p-value < 0.05.

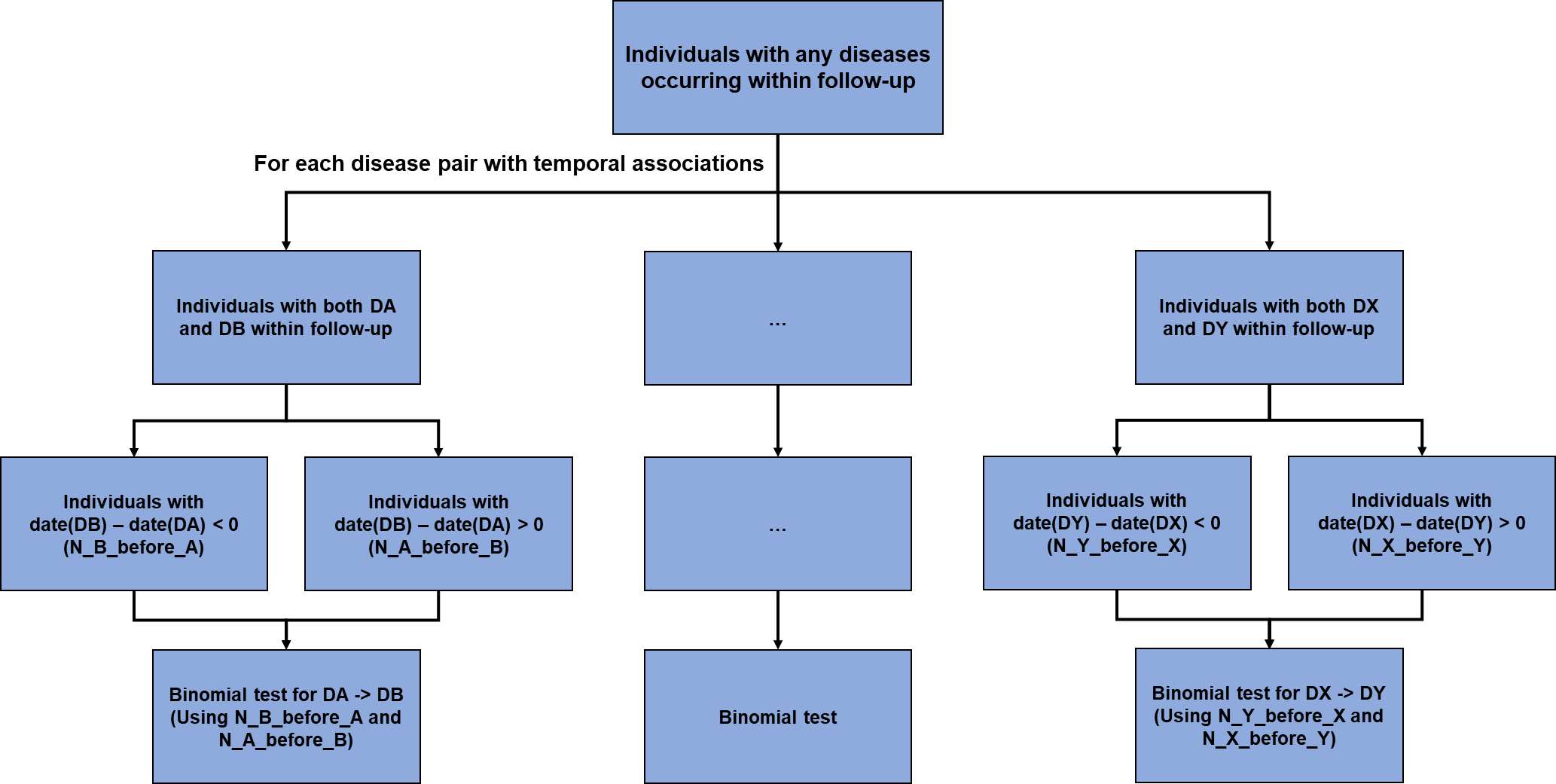

**Figure S4.** **Process for filtering individuals and diseases in the binomial test for determining disease pair directionality.**

***Conditional logistic regression***

For disease pairs with confirmed directionality (D1 → D2), individuals with D1 occurring outside the follow-up period are excluded. Next, individuals with D2 occurring before the follow-up period are also excluded. The remaining individuals are those with D2 occurring within the follow-up period, as well as those without D2 occurring. A case-control dataset is then generated, with D2 as the outcome and D1 as the exposure, using matching variables. Conditional logistic regression is then performed with D2 as the response variable and D1 as the explanatory variable (**Figure S5**). Disease pairs that form disease trajectories are those with a Bonferroni-adjusted p-value < 0.05 and a positive coefficient for D1.

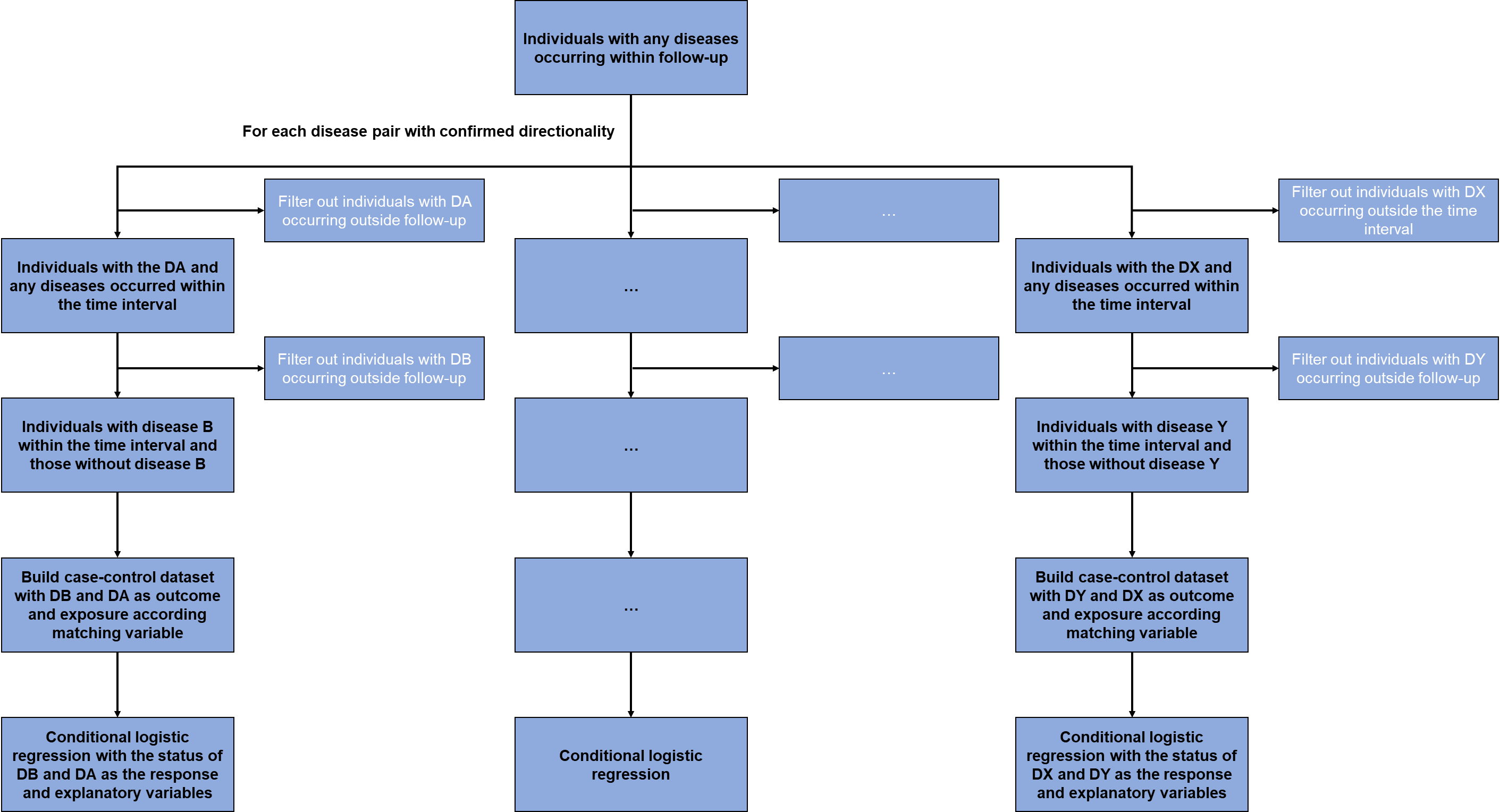

**Figure S5.** **Process for filtering individuals and diseases in conditional logistic regression.**

**LUKB DT Deployment and Usage**

**1. Deployment**

LUKB DT is an analysis component of LUKB, a tool designed for preparing UK Biobank data [1]. To use LUKB DT, LUKB must first be deployed. The deployment and access instructions for LUKB are detailed in the supplementary materials of the LUKB publication and are also available on the GitHub page: https://github.com/HaiGenBuShang/LUKB.

**2. Usage**

After logging into LUKB, select the “**Disease Trajectory Analysis**” tab to start building disease trajectories. The uploaded data file must contain at least three columns: individual ID, diagnoses, and corresponding diagnosis dates, separated by either a comma (,) or a tab (\t). Additional covariates may also be included. The data file can be uploaded by clicking the “**Browse**” button (**Figure S6 1**).

After uploading the file, a quick review of the uploaded data is provided to ensure correctness. Next, the analysis parameters should be set (**Figure S6 2**), including: 1) the specified exposure, 2) start and end dates for follow-up, 3) the minimum disease prevalence, 4) minimum Gap between two diseases, 5) Adjusting and matching variables for Cox regression and conditional logistic regression, and 6) the number of controls matched for each case. Note that the matching variables for conditional logistic regression should be categorical variable.

Once the parameters are set, click the “**Preview**” button (Figure **S6 3**) to generate a brief summary of the data. After reviewing the summary, click “**Confirm Parameters**” to save the data into LUKB (**Figure S6 4**).

The returned file prefix can then be used to start and track the following steps: 1) Cox regression (**Figure S6 5a**), 2) disease pair detection (**Figure S6 6a**), 3) conditional logistic regression (**Figure S6 7a**), and 4) construction of disease trajectories originating from the specified exposure (**Figure S6 8a**). Multiple computation cores can be specified for Cox regression and conditional logistic regression (**Figure S6 5a and 7a**).

The results of Cox regression, disease pair detection, conditional logistic regression, and disease trajectory construction are stored in **RData** format and can be downloaded by clicking the respective Download button (**Figure S6 5b, 6b, 7b, and 8b**).

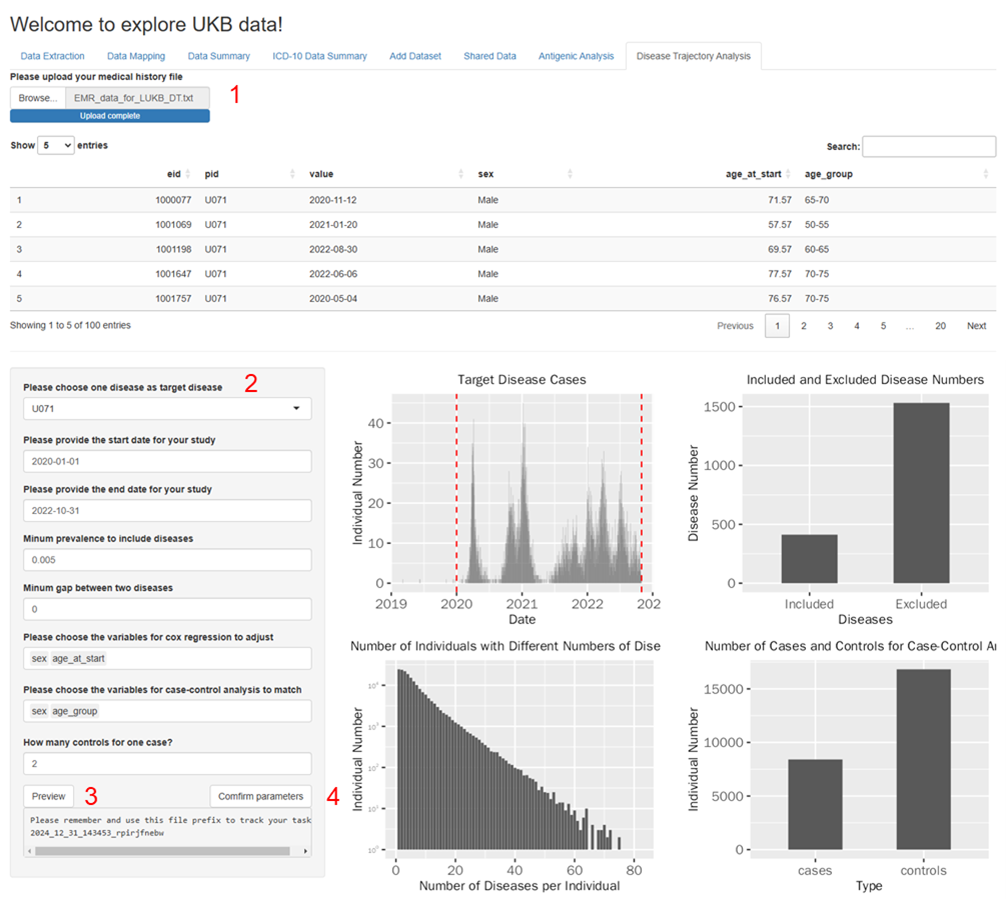

**Figure S6. LUKB DT usage.** 1) upload EMR data; 2) set analyzing parameters; 3) preview input data; 4) save data into LUKB; 5a) start and track Cox regression task; 5b) download Cox regression results; 6a) start and track disease pair detection task; 6b) download disease pair detection results; 7a) start and track conditional logistic regression task; 7b) download conditional logistic regression results; 8a) construct disease trajectories from the specified exposure; 8b) download disease trajectories.

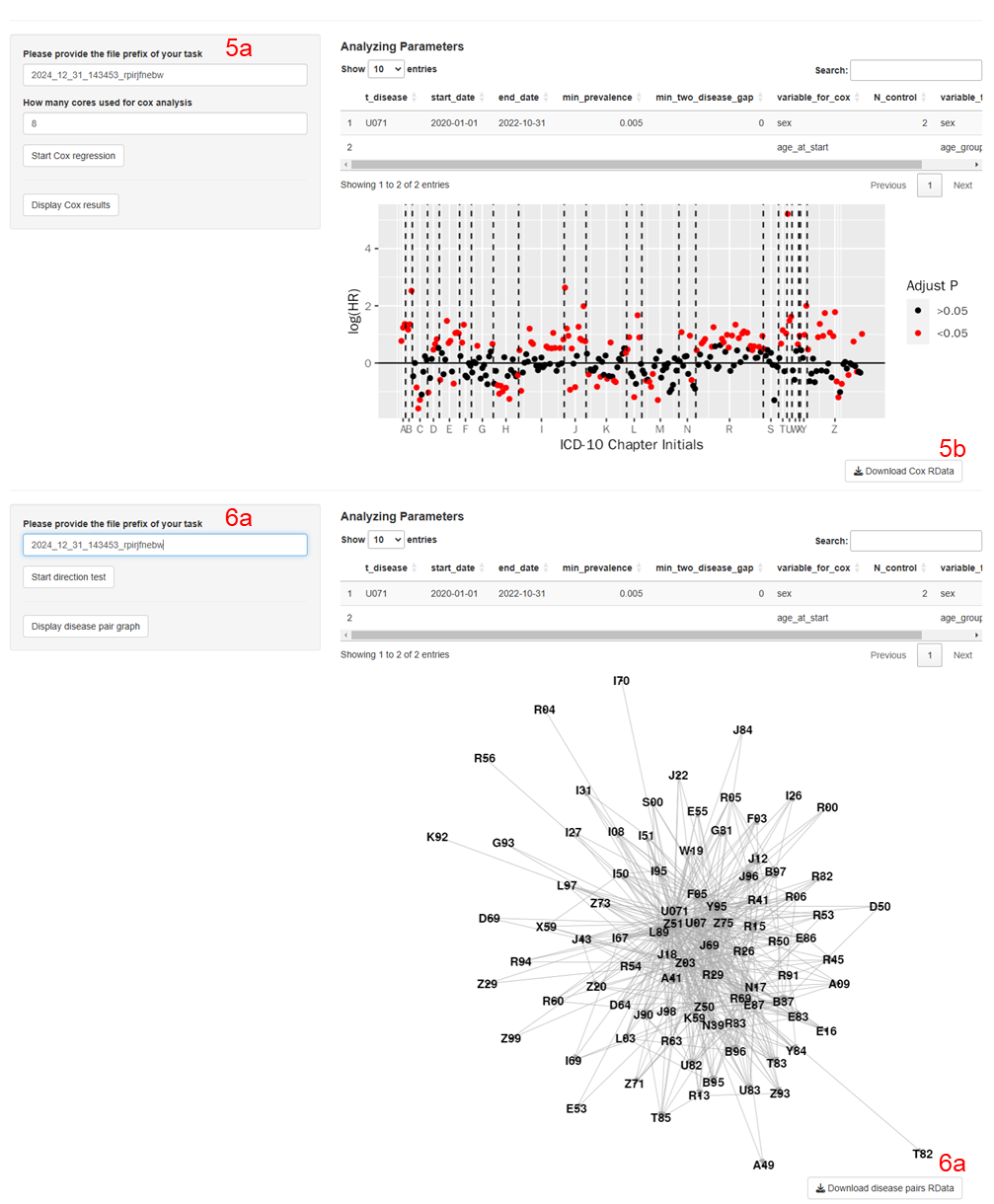

**Figure S6. LUKB DT usage (continued).** 1) upload EMR data; 2) set analyzing parameters; 3) preview input data; 4) save data into LUKB; 5a) start and track Cox regression task; 5b) download Cox regression results; 6a) start and track disease pair detection task; 6b) download disease pair detection results; 7a) start and track conditional logistic regression task; 7b) download conditional logistic regression results; 8a) construct disease trajectories from the specified exposure; 8b) download disease trajectories.

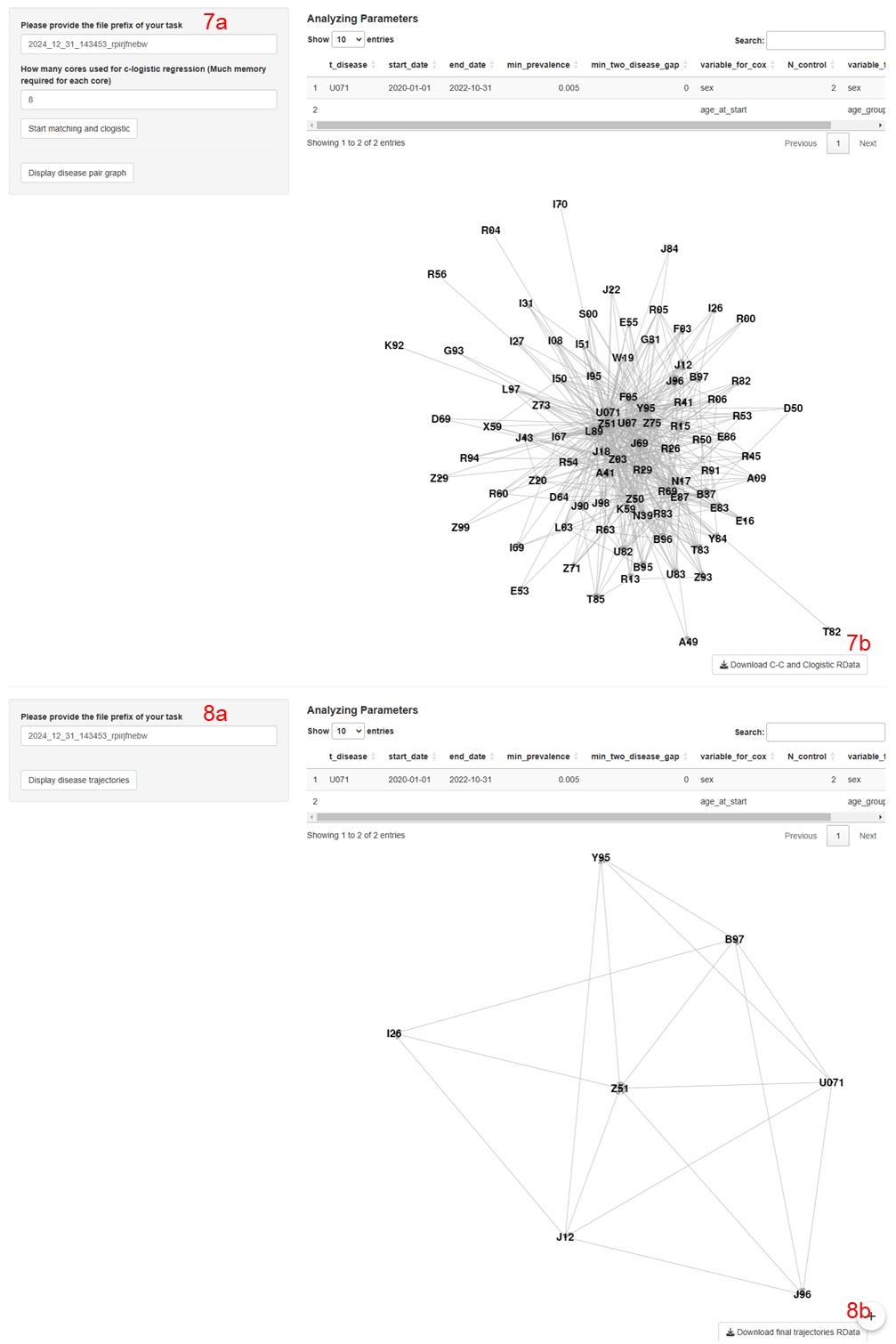

**Figure S6. LUKB DT usage (continued).** 1) upload EMR data; 2) set analyzing parameters; 3) preview input data; 4) save data into LUKB; 5a) start and track Cox regression task; 5b) download Cox regression results; 6a) start and track disease pair detection task; 6b) download disease pair detection results; 7a) start and track conditional logistic regression task; 7b) download conditional logistic regression results; 8a) construct disease trajectories from the specified exposure; 8b) download disease trajectories.
